# Type 2 diabetes and chronic gastritis/duodenitis comorbidity: additive risk for incident depression and synergistic risk for all-cause mortality – a retrospective cohort study

**DOI:** 10.1101/2025.08.17.25333765

**Authors:** Bo Hu, Yu-Ling Cui, Yan-Yu Li, Xin-Wen Yu, Hao Xie, Li-Juan Du, Shuo Guo, Yao Tong, Xiao-Yan Bai, Min-Hua Ni, Ai-Li Yang, Yu-Xin Jin, Sheng-Ru Liang, Lin-Feng Yan, Bin Gao, Guang-Bin Cui, Ying Yu

## Abstract

**Background:** Type 2 diabetes (T2D) and chronic gastritis/duodenitis (CGD) are both associated with the onset of depression and mortality. However, the impact of T2D-CGD comorbidity on incident depression and mortality remains unclear.

**Method:** This retrospective cohort study utilized data from 387,149 participants in the UK Biobank to examine the relationship between T2D-CGD comorbidity, incident depression, and all-cause mortality.

**Outcome:** Patients with T2D are more likely to develop CGD compared to those without T2D (odds ratio = 2·10, 95% CI = [1·97, 2·24]). T2D, CGD, and their comorbidity each independently increased risks of depression incidence and all-cause mortality, with T2D-CGD showing the strongest associations for both (depression incidence: adjusted hazard ratio [aHR] = 2·29, 95% CI = [1·84, 2·85]; all-cause mortality: aHR = 2·57, 95% CI = [2·28, 2·88]). The synergistic effect of T2D and CGD on all-cause mortality was 1·92 times that of their individual effects combined (synergy index = 1·92, 95% CI = [1·56, 2·31]). The comorbidity was associated with a higher risk of depression and all-cause mortality within 15 years of disease onset. White matter hyperintensity, particularly near the cerebral ventricles, partially mediated the relationship between T2D-CGD comorbidity and incident depression.

**Interpretation:** Integrated screening and long-term monitoring strategies should be prioritized for population with the comorbidity of T2D and CGD, as it significantly elevates the risk of both incident depression and all-cause mortality.

## 1. Introduction

Depression has emerged as a clinically significant comorbidity strongly linked to type 2 diabetes (T2D). Extensive evidence demonstrates that patients with T2D face a 1·5 to 2·0-fold increased risk of developing depression compared to non-diabetic individuals^1^. Moreover, both depression and subthreshold depressive symptoms have been consistently associated with higher risks of dementia onset and all-cause mortality^2,3^.

Chronic gastritis is a persistent gastric mucosal inflammation mainly caused by *H. pylori* infection, bile reflux, medication use, or alcohol consumption, leading to atrophy/metaplasia and potential dysplasia. Given the overlapping etiology, clinical manifestations, and therapeutic management of gastritis and duodenitis—as well as their anatomical continuity—both conditions are classified under the code K29 of the International Classification of Diseases, Tenth Revision (ICD-10)^4^. Chronic gastritis or duodenitis (CGD) is prevalent in type 2 diabetes (T2D) primarily due to high *H. pylori* susceptibility in T2D patients^5,6^, which worsens glycemic control and insulin resistance in reverse^7,8^. Growing evidence from translational research suggests that CGD may predispose to depression through gut-brain axis dysregulation ^9-11^. Despite these plausible connections, the interplay between T2D-CGD comorbidity, depression incidence, and all-cause mortality remains insufficiently explored.

This research utilized the data from the UK Biobank to conduct a retrospective cohort study, investigating the association between T2D-CGD comorbidity, incident depression, and all-cause mortality. Building on prior research that suggests the relationship between T2D, CGD, and depression risk may be mediated by microvascular dysfunction, neurodegenerative pathology, and neuroinflammation^1,12,13^—typically observed as white matter hyperintensities (WMH) on brain MRI—this study further explored the potential mediating role of WMH in the connection between T2D-CGD comorbidity and depression.

## 2. Methods

### 2.1 Study design, participants inclusion, and groups

This study utilized data from the UK Biobank (Approved Project ID: 627927), a large-scale, ongoing cohort study encompassing over 500,000 participants in the United Kingdom. Between 2006 and 2010, all participants underwent comprehensive baseline assessments^14^. Written informed consent was obtained from all participants, and ethical approval was granted by the North West Multicenter Research Ethics Committee. As the data were fully anonymized, no additional institutional review was required for this analysis. The study adhered to the Strengthening the Reporting of Observational Studies in Epidemiology (STROBE) guidelines.

This retrospective cohort study included 502,128 subjects. To minimize biases related to comorbid conditions, 14,069 participants with dementia, schizophrenia, bipolar disorder, brain tumors, stroke, type 1 diabetes, malnutrition-related diabetes, gestational diabetes, and acute gastritis were excluded. To ensure temporal precedence and reduce confounding, 100,910 individuals with baseline depression or those who developed T2D or CGD during the follow-up period were excluded. Finally, 387,149 participants were included in the final analysis (**Fig. 1**) and were divided into four groups: (1) Patients with neither T2D nor CGD (Control group); (2) Patients with T2D alone (T2D group); (3) Patients with CGD alone (CGD group); (4) Patients with T2D-CGD comorbidity (COM group).

**Fig. 1.**
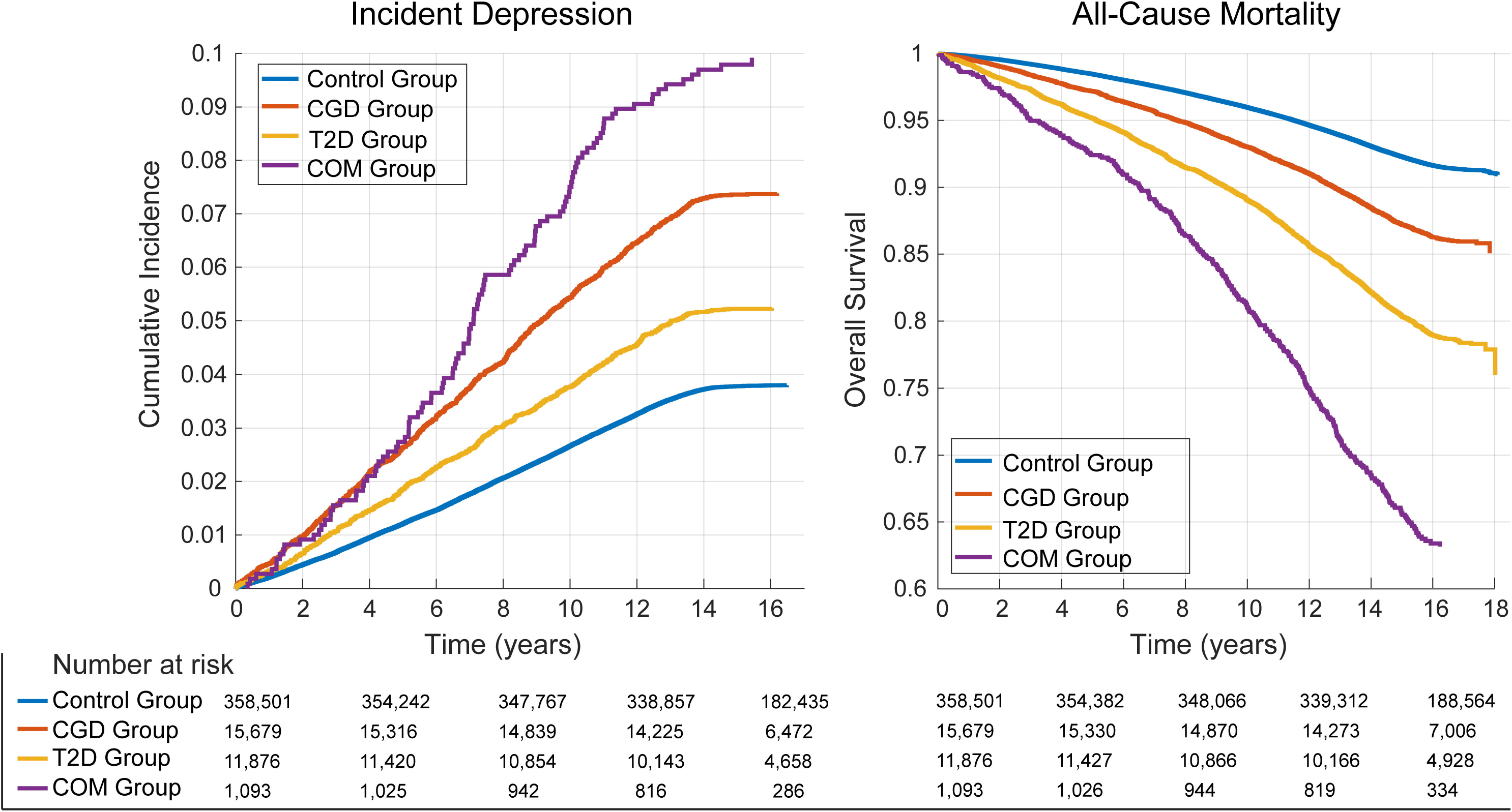
Flowchart depicting participant selection and subgrouping.

### 2.2 Definition of T2D, CGD, and depression

The UK Biobank captures disease onset dates through multiple linked data sources, including hospital episode statistics, primary care records, death register linkage, and others. Patients with T2D were identified using four methods: 1) ICD-10 codes E11 (non-insulin-dependent diabetes mellitus); 2) blood glucose ≥ 11·1 mmol/L at baseline; 3) HbA1c ≥ 48 mmol/mol at baseline; and 4) self-reported T2D diagnosis and dates. Patients with CGD were identified based on ICD-10 codes K29 (gastritis and duodenitis), excluding K29·0 (acute hemorrhagic gastritis) and K29·1 (other acute gastritis). Incident depression cases were identified using ICD-10 codes F32 (depressive episode) and F33 (recurrent depressive disorder).

### 2.4 Potential risk factors

A total of 18 potential risk factors were identified: 1) five disease-related factors, including T2D status, CGD status, comorbidity of both, T2D duration, and CGD duration; 2) three demographic factors, including age, sex (assigned at birth), and body mass index (BMI); 3) two environmental factors, including the Townsend deprivation index (TDI) and urbanization level (**supplementary appendix 1**); and 4) eight modifiable lifestyle factors, including sleep duration, fruit and vegetable intake, meat intake, bread and cereal intake, tea intake, coffee intake, smoking frequency, and alcohol consumption frequency (supplementary **appendix 2**).

### 2.5 Primary and secondary outcomes

The primary outcome of this study was incident depression, while the secondary outcome was all-cause mortality. Participants who did not reach either the primary or secondary outcome were right-censored at the date of March 31, 2025, which marked the latest update from the UK Biobank.

### 2.6 Imaging data

T2-weighted fluid-attenuated inversion recovery (FLAIR) data were acquired using a Siemens Skyra 3T scanner with a standard Siemens 32-channel head coil. Two indicators of WMH were used: 1) Peri-ventricular WMH (pvWMH), which are located within 10 mm from the lateral ventricle wall, primarily involving the radiations of the corpus callosum and the medial part of the centrum semiovale; 2) Deep WMH (deepWMH), situated more than 10 mm away from the ventricle in deep white matter regions, typically in the frontal, parietal, temporal, and occipital lobes, as well as the corona radiata above the basal ganglia.

### 2.7 Statistical analysis

All statistical analyses were performed using in-house codes on the MATLAB R2018a platform (MathWorks, Natick, MA). Baseline demographic characteristics were summarized as percentages for categorical variables and means with standard deviations (SDs) for continuous variables. Participants with missing data were excluded from corresponding analyses. The odds ratio (OR) was calculated to quantify the association between T2D and CGD. Kaplan-Meier (K-M) survival curves were used to estimate cumulative incidence of depression and all-cause mortality across groups. Multivariable Cox regression analyses assessed the risks of incident depression and all-cause mortality linked to the comorbidity of T2D and CGD, compared to the control group. Five models adjusted for different confounders were used: model 1 (unadjusted), model 2 (T2D and CGD status and durations), model 3 (model 2 plus age, sex, and BMI), model 4 (model 3 plus TDI and urbanization level), and model 5 (model 4 plus eight modifiable lifestyle factors). The Fine and Gray method accounted for competing risks of death. Hazard ratios (HRs) with 95% confidence intervals (CIs) were reported to assess statistical significance. The proportional hazard assumption was tested using Schoenfeld residuals, which were satisfied.

Relative excess risk due to interaction (RERI), attributable proportion due to interaction (AP), and synergy index (SI) ^15^ were used to determine whether the combined effect of T2D and CGD on the risk of incident depression and all-cause mortality exceeded the simple additive effects of the individual conditions (**supplementary appendix 3**). The bootstrap method with 10,000 resamples was employed to estimate 95% CIs for these three indicators.

To further explore underlying mechanisms, cerebral WMH was used for mediation analysis in the relationship between T2D-CGD comorbidity and incident depression, adjusting for age, sex, and BMI as covariates. The indirect effect (mediation effect) was calculated, and the bootstrap method with 10,000 resamples was applied to estimate 95% CIs.

## 3. Results

### 3.1 Demographic information and the association between T2D and CGD

The demographic information is summarized in **Table 1**. A significant positive association was found between T2D and CGD, with an OR of 2·10 (95% CI: [1·97, 2·24]), indicating that individuals with T2D had 2·1 times the odds of developing CGD compared to those without T2D.

### 3.2 K-M analysis and multivariable Cox regression analysis of incident depression

The K-M survival curves in **Fig. 2** illustrate that COM group exhibited the highest incidence of depression, followed by the CGD group, T2D group, and Control group (log-rank test P < 0·0001). Multivariable Cox regression analysis revealed that T2D-CGD comorbidity was significantly associated with a higher risk of incident depression across five models: model 1 (univariable; HR = 2·56, 95% CI = [2·12, 3·09]), model 2 (adjusted for T2D and CGD status and duration; aHR = 2·90, 95% CI = [2·37, 3·58]), model 3 (model 2 plus age, sex, and BMI; aHR = 2·76, 95% CI = [2·24, 3·41]), model 4 (model 3 plus TDI and urbanization level; aHR = 2·48, 95% CI = [2·00, 3·06]), and model 5 (model 4 plus eight modifiable lifestyle factors; aHR = 2·29, 95% CI = [1·84, 2·85]). In addition, both T2D and CGD were independently associated with a higher risk of incident depression in model 5 (T2D: aHR = 1·19, 95% CI = [1·08, 1·32]; CGD: aHR = 2·00, 95% CI = [1·83, 2·20]). All aHRs and 95% CIs for model 5 are presented in **Fig. 3** (**supplementary Fig. 1** for other models). The Fine and Gray method was applied for competing risk analysis to account for bias from competing risks of death, resulting in an aHR for model 5 of 1·95 (95% CI = [1·48, 2·56]).

**Fig. 2.** Kaplan-Meier analysis comparing depression incidence and all-cause mortality among patients with T2D-CGD comorbidity (COM group), T2D alone (T2D group), CGD alone (CGD group), and no conditions (Control group).

**Fig. 3.** Effect of T2D-CGD comorbidity on depression incidence and all-cause mortality after adjusting for clinical, demographic, environmental, and modifiable lifestyle risk factors. Red box: potential risk factors (HR>1); Blue box: potential protective factors (HR<1).

### 3.3 K-M analysis and multivariable Cox regression analysis of all-cause mortality

The K-M survival curves in **Fig. 2** demonstrate that the COM group had the highest incidence of all-cause mortality, followed by the T2D group, CGD group, and Control group (log-rank test P < 0·0001). Cox regression analysis revealed that T2D-CGD comorbidity was significantly associated with a higher risk of all-cause mortality across five models: model 1 (HR = 4·81, 95% CI = [4·36, 5·31]), model 2 (aHR = 5·21, 95% CI = [4·65, 5·83]), model 3 (aHR = 2·90, 95% CI = [2·58, 3·25]), model 4 (aHR = 2·67, 95% CI = [2·38, 2·99]), and model 5 (aHR = 2·57, 95% CI = [2·28, 2·88]). In addition, both T2D and CGD were independently associated with a higher risk of all-cause mortality in model 5 (T2D: aHR = 1·68, 95% CI = [1·59, 1·77]; CGD: aHR = 1·28, 95% CI = [1·19, 1·36]). All aHRs and 95% CIs for model 5 are displayed in **Fig. 3** (**supplementary Fig. 2** for other models).

### 3.4 Analysis of synergistic effect

The synergistic effect of T2D and CGD on depression risk did not exceed the sum of their individual effects (RERI = 0·38, 95% CI = [-0·20, 1·04]; AP = 0·13, 95% CI = [-0·09, 0·30]; SI = 1·26, 95% CI = [0·86, 1·72]). In contrast, the synergistic effect of T2D and CGD on all-cause mortality surpassed the sum of their individual effects (RERI = 1·23, 95% CI = [0·76, 1·76]; AP = 0·34, 95% CI = [0·24, 0·43]; SI = 1·92, 95% CI = [1·56, 2·31]). This result suggests that the combined effect of T2D and CGD on all-cause mortality is 1·92 times greater than the sum of their individual effects.

### 3.5 Stratified analysis by follow-up duration

When the follow-up time was stratified as < 5 years, 5–10 years, and >10 years, the adjusted HRs (aHRs) and 95% CIs for the association between T2D-CGD comorbidity and depression (model 5) were as follows: aHR = 2·34 (95% CI = [1·57, 3·50]), aHR = 2·54 (95% CI = [1·83, 3·53]), and aHR = 1·22 (95% CI = [0·80, 1·86]) for each time interval, respectively (**supplementary Fig. 3**). The aHRs and 95% CIs for the association between T2D-CGD comorbidity and all-cause mortality (model 5) were: aHR = 3·59 (95% CI = [2·79, 4·63]), aHR = 2·86 (95% CI = [2·32, 3·52]), and aHR = 1·20 (95% CI = [1·01, 1·42]), respectively (**supplementary Fig. 4**).

In summary, T2D-CGD comorbidity was linked to the risk of depression and all-cause mortality within 15 years following disease onset (with baseline disease durations of approximately 5 years). Depression incidence initially increased, then decreased, while all-cause mortality displayed a gradual decline.

### 3.5 Stratified analysis by sex

When we manually divided all participants into two group according to sex, the aHRs and 95% CIs for the association between T2D-CGD comorbidity and depression (model 5) were aHR = 2·26 (95% CI = [1·64–3·13]) in female group, and aHR = 2·29 (95% CI = [1·71–3·08]) in male group (**supplementary Fig. 5**). The aHRs and 95% CIs for the association between T2D-CGD comorbidity and all-cause mortality (model 5) were aHR = 2·67 (95% CI = [2·15–3·31]) in female group, and aHR = 2·48 (95% CI = [2·16–2·85]) in male group (**supplementary Fig. 6**)

In brief, the risk of T2D-CGD comorbidity for depression and all-cause mortality was similar between female and male groups.

### 3.6 Mediation analysis

Mediation analyses were conducted to explore whether cerebral WMH volume mediated the association between T2D-CGD comorbidity and incident depression. A total of 54,060 participants who underwent MRI scans were included in this analysis (**supplementary appendix 4**). The mediation analysis revealed significant indirect effects *via* both WMH subtypes: pvWMH showed an indirect effect of 0·019 (95% CI: 0·008–0·032), while deepWMH demonstrated an indirect effect of 0·007 (95% CI: 0·001–0·014), indicating that the association between T2D-CGD comorbidity and incident depression was partially mediated by WMH, particularly pvWMH (**supplementary Fig. 7**).

### 3.7 Sensitive analysis

To assess the robustness of these findings, this study tested a scenario where only diagnostic data (ICD-10 codes) were used to define T2D, excluding participants based on other criteria (blood biochemical data and self-reported diagnosis). This analysis included 9,101 participants (76·6%) and yielded results consistent with the original findings (**supplementary Fig. 8**).

Additionally, depression was defined to include both depressive episodes and recurrent depressive disorders. By including only participants with depressive episodes (15,474 participants, 98·7%) and excluding others, the results remained consistent with the original findings (**supplementary Fig. 9**).

## 4. Discussion

Few studies have examined the impact of T2D-CGD comorbidity on depression incidence and all-cause mortality. This cohort study found that patients with T2D are more likely to develop CGD compared to those without T2D. T2D, CGD, and their comorbidity each independently increased risks of depression incidence and all-cause mortality, with their comorbidity showing the strongest associations for both depression incidence and all-cause mortality. The synergistic effect of T2D and CGD on all-cause mortality was 1·92 times that of their individual effects combined. The T2D-CGD comorbidity was associated with an increased risk of depression and all-cause mortality within 15 years of disease onset. Mediation analysis revealed that WMH, particularly those near the cerebral ventricle, partially mediated the association between T2D-CGD comorbidity and incident depression.

The relationship between T2D and CGD have been confirmed in previous genetical research^6^, substantial evidence also links T2D to *H. pylori* infection^5,8,16^. Most studies report OR or relative risks (RR) ranging from 1·08 to 2·00 for CGD or *H. pylori* infection in patients with T2D, although some findings contradict this association^17^ or even suggest a protective effect of *H. pylori* against T2D^18^. These inconsistencies may result from population heterogeneity or unresolved causal mechanisms. While *H. pylori*-induced insulin resistance, chronic inflammation, and metabolic syndrome are widely considered drivers of T2D, other studies indicate that hyperglycemia in patients with T2D may impair *H. pylori* eradication efficacy^19^. Furthermore, dietary irregularities associated with T2D could also exacerbate CGD incidence. These findings suggest a bidirectional, multifactorial interaction between T2D and CGD rather than a unidirectional causal link.

The association between T2D and depression has been well established^20^, with shared pathogenic mechanisms including genetic predisposition, insulin resistance, microvascular damage, chronic inflammation, and gut-brain axis dysregulation^12,21,22^. Notably, while CGD is more strongly associated with depression than T2D in this study, there is fewer research investigated the relationship between CGD and depression than T2D^11^. Most existing studies focus on gut-brain interaction disorders^9,10,23^, which only partially overlap with CGD and require distinct therapeutic approaches. Regarding mortality, *H. pylori* infection synergistically increases all-cause mortality in patients with T2D^24,25^. Importantly, CGD and *H. pylori* infection are distinct yet overlapping conditions—not all patients with CGD are *H. pylori*-positive, nor do all *H. pylori* carriers develop CGD. This distinction underscores the critical need for direct investigation into T2D-CGD comorbidity to guide evidence-based clinical management strategies.

The association between WMH and depression observed in our study is consistent with established neuroimaging evidence^26,27^. Diffusion MRI studies consistently demonstrate a correlation between depression and impaired white matter microstructural integrity^28^. Our findings specifically highlight the mediating role of pvWMH in the relationship between T2D-CGD comorbidity and depression, supporting the results of previous studies^27^. Mechanistically, T2D-CGD comorbidity contributes to depression may *via* cerebrovascular dysfunction characterized by: (1) endothelial cell damage impairing the blood-brain barrier, and (2) venous outflow obstruction resulting in interstitial fluid accumulation and periventricular edema^29^. Notably, this impairment in cerebrospinal fluid-interstitial fluid exchange shares pathophysiological features with Alzheimer’s disease, suggesting potential overlapping neurodegenerative mechanisms in these comorbid conditions^30^.

Several limitations must be considered. As noted in the discussion section, the causal relationships between T2D, CGD, and depression remain complex and unresolved in this observational study. Further mechanistic research is required to clarify these relationships. Additionally, some factors may not have been accounted for, potentially confounding or driving the associations observed. For example, the regularity of dietary patterns and medication use over extended periods cannot be analyzed in the current sample. Selection bias may also be a concern in the UK Biobank, primarily due to its reliance on hospital data, primary care records, and death registration. Thus, patients who were depressed but did not seek medical treatment may have been missed, potentially underestimating the effect size of the exposure association or delaying diagnosis.

## Conclusion

Integrated screening and long-term monitoring strategies should be prioritized for population with the comorbidity of T2D and CGD, as it significantly elevates the risk of both incident depression and all-cause mortality.

## Data Availability

All data produced are available online at UK Biobank.

https://www.ukbiobank.ac.uk/

## Author Contributions

Bo Hu, Yan-Yu Li, Hao Xie: Conceptualization, Methodology, Formal Analysis, Investigation, Accessed and Verified the Data, Writing – Original Draft, Writing – Review & Editing.

Xin-Wen Yu, Ai-Li Yang, Yu-Xin Jin, Sheng-Ru Liang: Conceptualization (Clinical Expertise), Validation, Resources, Writing – Review & Editing.

Li-Juan Du, Shuo Guo, Yao Tong, Xiao-Yan Bai: Accessed and Verified the Data, Formal Analysis (partial), Visualization. Min-Hua Ni, Tian Chai, Lin-Feng Yan: Accessed and Verified the Data, Validation Analysis.

Bin Gao, Guang-Bin Cui, and Ying Yu: Accessed and Verified the Data, Project Administration, Supervision, Formal Analysis (Oversight), Writing – Original Draft, Writing – Review & Editing (Revision).

## Declare of interests

All authors declare no competing interests.

## Data sharing

All data are available from the UK Biobank.

## Acknowledgments

BH has received funding from the National Natural Science Foundation of China [grant number 82302148]. YY has received funding from the Key Science and Technology Program of Shaanxi Province [grant number 2023-YBSF-331] and Fourth Military Medical University [grant number 2023XC045]. GBC has received funding from the National Natural Science Foundation of China [grant number 82471936].

